# Statistical tests for heterogeneity of clusters and composite endpoints

**DOI:** 10.1101/2021.06.16.21258900

**Authors:** Anthony J. Webster

## Abstract

Clinical trials and epidemiological cohort studies often group similar diseases together into a composite endpoint, to increase statistical power. A common example is to use a 3-digit code from the International Classification of Diseases (ICD), to represent a collection of several 4-digit coded diseases. More recently, data-driven studies are using associations with risk factors to cluster diseases, leading this article to reconsider the assumptions needed to study a composite endpoint of several potentially distinct diseases. An important assumption is that the (possibly multivariate) associations are the same for all diseases in a composite endpoint (not heterogeneous). Therefore, multivariate measures of heterogeneity from meta-analysis are considered, including multi-variate versions of the *I*^2^ and *Q* statistics. Whereas meta-analysis offers tools to test heterogeneity of clustering studies, clustering models suggest an alternative heterogeneity test, of whether the data are better described by one, or more, clusters of elements with the same mean. The assumptions needed to model composite endpoints with a proportional hazards model are also considered. It is found that the model can fail if one or more diseases in the composite endpoint have different associations. Tests of the proportional hazards assumption can help identify when this occurs. It is emphasised that in multi-stage diseases such as cancer, some germline genetic variants can strongly modify the baseline hazard function and cannot be adjusted for, but must instead be used to stratify the data.

## Introduction

It is common for epidemiological studies [1] and clinical trials [2–4] to group similar diseases together into a cluster of diseases, providing more total cases, and more statistical power to detect associations. This procedure has been essential since the pioneering epidemiological studies of John Graunt in the 1600s [5], providing sufficient cases to allow meaningful statistical study, while attempting to ensure that clustered diseases have a similar etiology. Today an increasingly detailed biological understanding informs the clustering of diseases. One common approach is to use the heirarchical classification of the International Classification of Diseases (ICD) as a guide for which diseases to group [6]. The ICD system clusters increasingly detailed disease descriptions into larger clusters with similar disease etiology, often allowing larger clusters to be used in epidemiological studies. In practice, diseases are selected by a clinician to help ensure that only diseases with a clearly defined and common etiology are clustered together [1]. With data-driven clustering studies increasingly being used to identify potential composite endpoints [1, 7–11], it seems appropriate to review the assumptions needed to study a cluster of diseases, and the existing arguments for and against doing so in clinical trials.

In composite endpoints, or diseases clustered using shared risk factors [1], a necessary requirement is that risk-factor associations are the same (homogeneous). A similar requirement arose in meta-analysis, and led to the development of heterogeneity tests [12]. The multivariate versions of these tests, their origin, and their application to assess the heterogeneity of risk-factor associations within a composite endpoint or a clustering study is described. In the process, an alternative clustering-based heterogeneity test is suggested that offers a different perspective to the conventional *Q* and *I*^2^ statistics [13, 14], that are commonly used in meta-analyses [12].

The proportional hazards modelling is ubiquitous in medical research and statistical epidemiology, so the implicit assumptions needed when using it to model a composite endpoint are explored. Heterogeneity of disease associations with risk-factors in a composite endpoint, are sufficient to cause the proportional hazards assumptions to fail. Therefore it is necessary for tests of the proportional hazards assumption to be satisfied, and these can be used when tests of heterogeneity are inappropriate due to insufficient data for example. From a statistical modelling perspective, the key requirement is that risk-factor associations are the same for diseases within a composite endpoint, baseline incidence rates can be arbitrarily different. (Although as will be discussed in the context of clinical trials, insufficient cases of a disease that prevent tests of heterogeneity, should only be included if there are strong prior reasons to expect the same risk-factor associations as for the other diseases in the endpoint.) The conceptual multi-stage model of diseases [15] such as cancer [16–18] and motor neuron disease [19–21], is used to highlight some additional implicit assumptions that are needed when the proportional hazards methodology is used.

### Composite endpoints in clinical trials

It is worth briefly recapping the benefits and concerns about using composite endpoints in clinical trials.

### Reasons to consider composite endpoints

There are several good reasons to use composite endpoints:

i. Diseases are implicitly defined as a composite endpoint of symptoms and biological measurements. (More precise sub-groups sometimes have different behaviours [9, 10, 22, 23].)
ii. Statistical power - If a drug targets a pathway that modifies the risk of several otherwise distinct diseases, or if a disease is a symptom of onset of more serious disease, then it makes sense to study these together to increase statistical power. The importance of both i) and ii) are apparent in the tables of disease studied by John Graunt in the 1600s [5], where it is difficult to compare diseases when cases are rare or erratic. Weaker arguments to use composite endpoints include:
iii. Interest is in avoiding any negative disease outcomes, not just particularly severe ones.
iv. Interest is in testing the potential influence of a potential drug or risk factor on a wide range of diseases.

These arguments iii) and iv) are more easily criticised as “fishing”, casting a wider net to try and catch more diseases that may be influenced by a new drug without strong prior reason to do so, and increasing the risk of false positive associations.

### Criticisms of composite endpoints

There are several serious criticisms and limitations of using composite endpoints, some of which are easily corrected, but others that appear unavoidable:

a. Early criticisms of composite endpoints were, to quote Cordoba et al. [24], that “Components are often unreasonably combined, inconsistently defined, and inadequately reported.”.
b. Endpoints can be of different importance to patients. This has led several authors to argue for weighting of endpoints based on importance to patients [2, 25, 26].
c. Infrequent disease - Insufficient cases of a disease in a composite endpoint can make it impossible to test if effect sizes are comparable, e.g. due to unreliable confidence intervals.
d. Similar effect sizes - If effect sizes are dissimilar, then it is impractical to meaningfully interpret results. If disease risk is being modified through a shared disease pathway, then we might expect a similar proportional reduction in disease risk.

Criticisms in a) can be avoided by careful study design, specification, and reporting. The concerns in c) limit studies to diseases with sufficient cases of each distinct disease to allow meaningful tests of similar effect sizes. A more nuanced concern is b), the argument that endpoints should be of equal importance to different patients. To develop understanding and treatments, it is most important to identify endpoints with a shared disease pathway. Whereas diseases sharing this pathway could have very different relevance to patients, diseases should only be included if they have the same underlying cause.

Despite the limitations, points (i) and (ii) emphasise that composite endpoints are often reasonable, or necessary, and in these cases it is important to understand the assumptions being made when we study them. Research that is intended to cluster diseases by common disease pathways will also lead to newly hypothesised composite endpoints, and it is helpful to establish tests and understand the assumptions that disease clusters are consistent with.

### Heterogeneity of associations

Heterogeneity tests are widely used in meta-analyses, and are intended to assess whether the reported associations are the same in several different studies [12]. For composite endpoints we wish to assess whether one or more associations are the same for all diseases in a composite endpoint. The method’s multivariate generalisation that is needed for clustering studies such as Ref. [1], is described below. Multivariate heterogeneity tests were originally developed for meta-analyses [27–29], for which case a random effects model is often more suitable. A discussion of the multivariate test for meta-analyses in a random effects model, can be found in [30]. Here we explain the basis for the (fixed effects) multivariate heterogeneity test, that is most relevant to composite endpoints. Later a test that originates from clustering studies is suggested, that offers an alternative approach with some advantages over conventional *Q* and *I*^2^ statistics [12].

The null hypothesis (in a fixed effects model), is that all diseases in a composite endpoint have the same associations with one or more parameters, such as a drug, or a collection of potential risk factors. These might be a subset of associations, with potential confounders adjusted for, and subsequently removed by marginalisation [1]. Consider *m* composite endpoints (or clusters of diseases), labelled by *g*. Under the null hypothesis of the same associations for diseases in a composite endpoint, labelled *i* = 1 to *i* = *n*_*g*_,

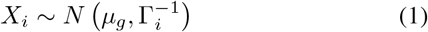

where 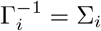 is the covariance (Γ_*i*_ is the precision matrix), and *µ*_*g*_ are the (unknown) associations that we are estimating, that are assumed to be the same for all diseases in the composite endpoint. Eq. 1 requires [31],

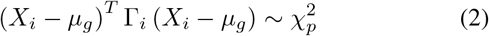

where *p* is the dimension. Therefore because the sum of *n*_*g*_ random variables that are individually 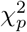 distributed is 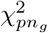,

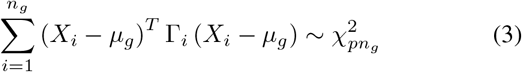

For *p* = 1 this has,

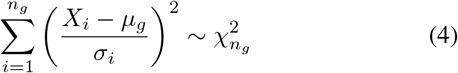

for standard deviations *σ*_*i*_. Because *µ*_*g*_ is unknown and must be estimated, the test statistic is modified, as explained next.

Using Bayes theorem with either a flat or normal prior for the mean *µ*_*g*_, a cluster of diseases with covariances {Γ_*i*_} and the same mean *µ*_*g*_, have [32],

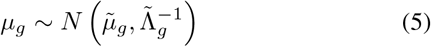

where,

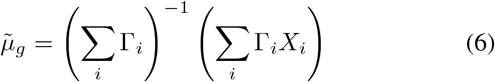

and,

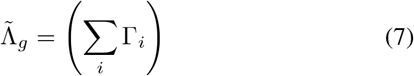

where if a normal prior is used then the sum over *i* includes the prior’s mean *µ*_0_ and covariance Λ_0_, and the sum is from *i* = 0 to *i* = *n*_*g*_. For a flat prior, the sum is from *i* = 1 to *i* = *n*_*g*_. The subscripts *g* allow the discussion to include more than one cluster of diseases, as was considered in Webster et al. [1]. Here unless stated otherwise, we consider a single composite endpoint, and the subscript *g* could be omitted.

As a result of Eq. 5, we have,

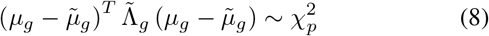

These observations, together with Eq. 1, can be used to derive a multivariate test for the assumption that the normal distributions have the same mean. Appendix A shows that,

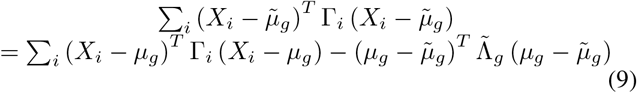

Using this with Eqs. 3 and 8,

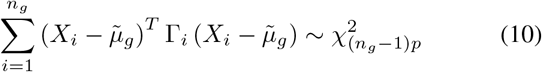

The left side of Eq. 10 is the Q statistic. It provides a test for the assumption that the (approximately) normally distributed estimates {*X*_*i*_} have the same mean, as is assumed for a composite endpoint or cluster of diseases. For *p* = 1, these expressions give the well known inverse variance weighted heterogeneity test, that are regularly used in meta analyses and 2-sample Mendelian randomisation studies [12, 33].

For the situation described in Webster et al. [1], the aim is to assess the goodness of fit for a clustering of diseases. In effect, there is a set of composite endpoints being considered, and the task is to determine the optimum split of diseases into composite endpoints with similar risk-factor associations. For this situation, Eq. 10 is modified to sum over all *m* clusters, and the Q statistic becomes,

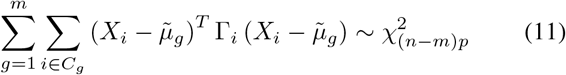

where we used 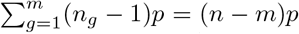, diseases in cluster and *C*_*g*_ is the set of diseases in cluster *g* (composite endpoint g).

The results above correspond to a fixed effects model where the data are assumed to have the same mean, as opposed to the means being sampled from an underlying distribution (a random effects model). A random effects model samples each study’s mean *µ*_*i*_, with 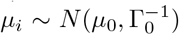, and assumes measured estimates *X*_*i*_ have 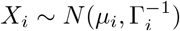 Marginalising out *µ*_*i*_ gives 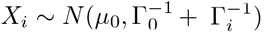, that replaces Eq. 1. Assuming that the data belong to a single cluster, then Eq. 5 becomes 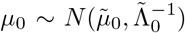, and Eqs. 6 and 7 become,

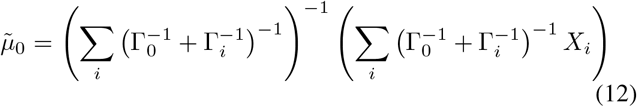

and

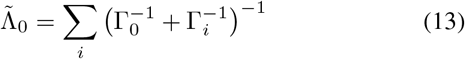

as in Jackson et al. [30] (where 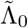 is the inverse of the covariance). To calculate the *Q* and *I*^2^ statistics using Eq. 10, Γ_*i*_ is replaced by 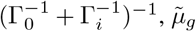 with 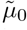, and *n*_*g*_ is the total number of studies. The above arguments and results could be modified to consider a random effects model with different priors for each cluster.

### Assessing heterogeneity within meta-analyses

The classical measure of heterogeneity uses the Q statistic, that was derived above in a multi-variate context. The I-square statistic [13, 14] is closely related to the Q statistic [12], and in the notation above, is for Eq. 11,

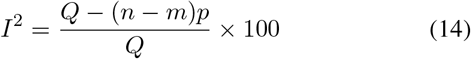

where *Q* is the left side of Eq. 11, and the factor of 100 is conventionally used to express *I*^2^ as a percentage. Unlike in figures 1 and 2, *I*^2^ is usually set to zero if its evaluation is negative. The equivalent expression for a single cluster that uses the left side of Eq. 10 for *Q*, would replace the number of degrees of freedom (*n − m*)*p*, with (*n*_*g*_ − 1)*p* in Eq. 14. The *I*^2^ statistic replaces a test with a more nuanced measure of heterogeneity that is particularly useful when some heterogeneity is expected, but it no longer provides an objective statistical test.

**Figure 1:**
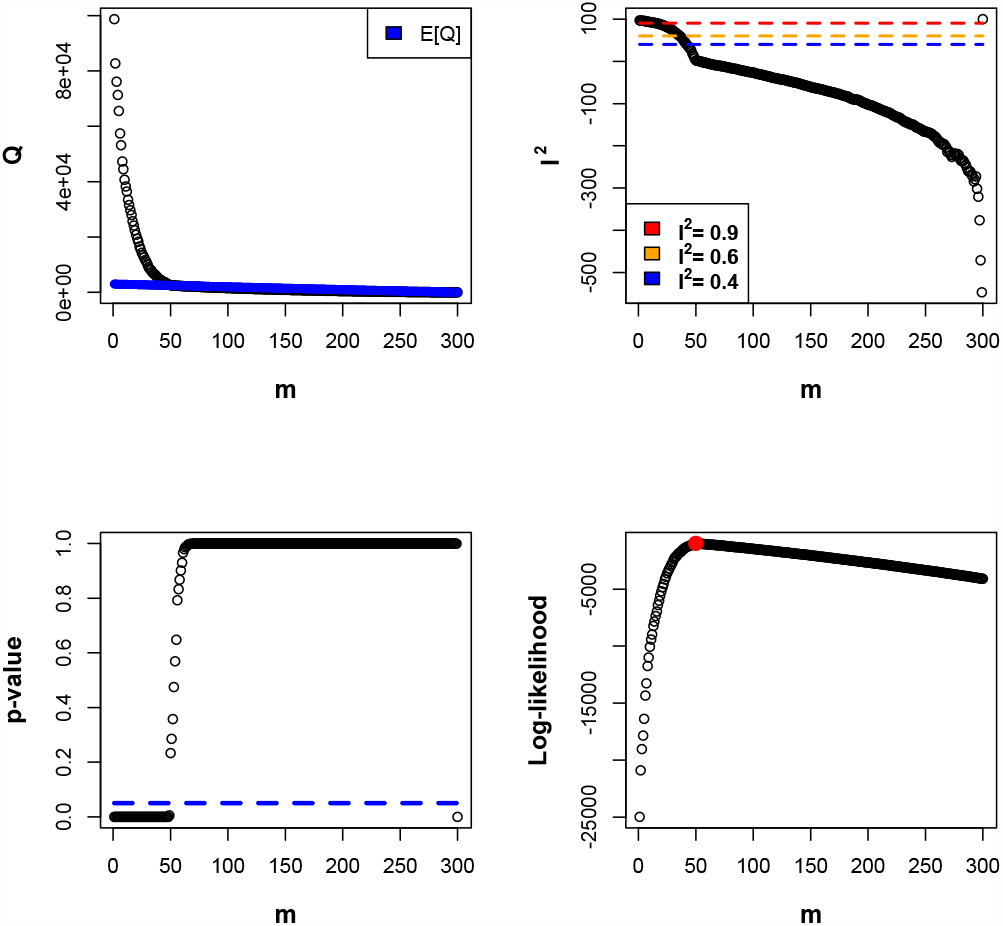
For a flat prior and normally distributed test data [32], the *Q*-statistic and log-likelihood correctly identify the 50 clusters of test data with 2 to 14 members each [32]. Interestingly, the rate of reduction of *I*^2^ slowed after the 50 clusters were identified.

**Figure 2:**
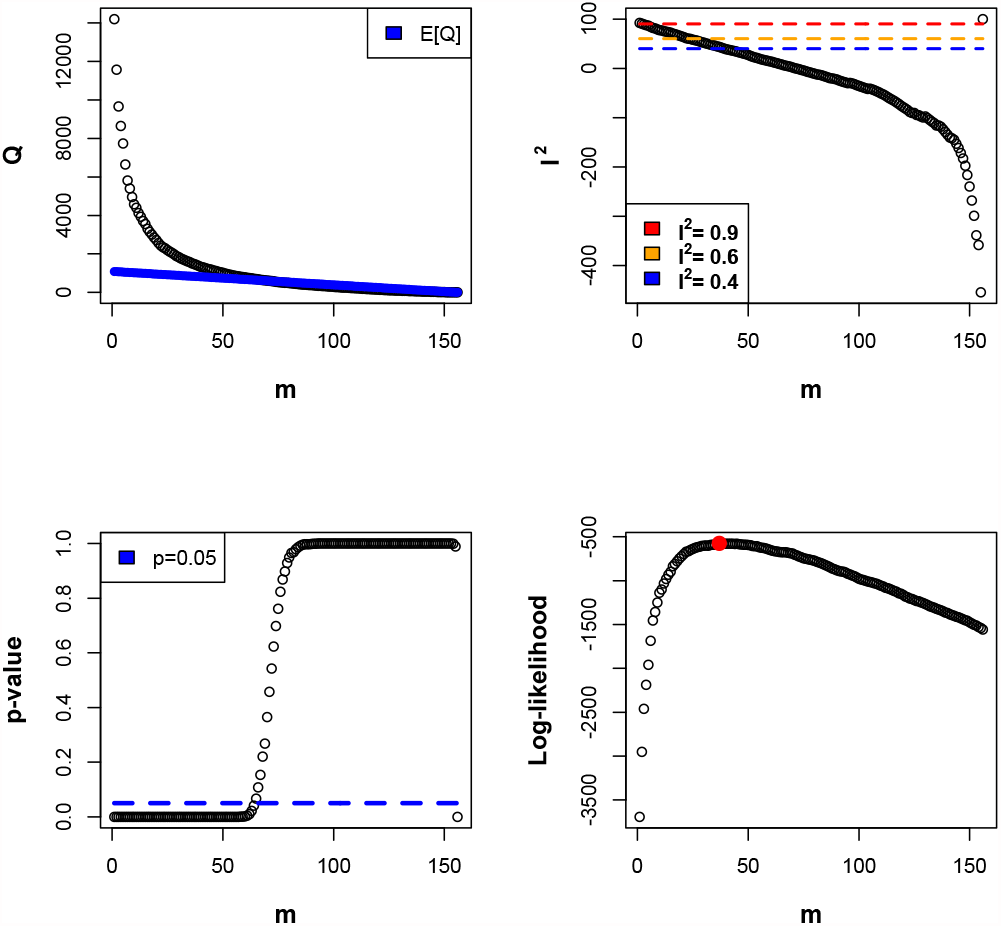
For a flat prior and UK Biobank disease data [1], the clustering log-likelihood of Webster [32] is minimised at *m* = 37, where *I*^2^ ≃ 50% A statistical heterogeneity test using *Q* fails for less than 63 clusters. With a normal prior centred on zero, *I*^2^ reduces and the maximum in log-likelihood moves to *m* ∼ 25, and was fairly insensitive to the prior’s covariance [32].

This article was originally motivated by the need to better understand and characterise the results from clustering studies, such as assessing the heterogeneity of the underlying clusters. However, clustering studies are becoming increasingly rigorous (Figures 1 and 2), and can offer an alternative approach to test the heterogeneity of composite endpoints. Webster [32] provides a log-likelihood for clustering normally distributed data such as that from maximum likelihood estimates, that assumes equal means in each cluster. The log-likelihood is maximised to determine whether the data fit more naturally in one or more clusters. This provides an intuitive and objective assessment of heterogeneity, that in the examples shown in figures 1 and 2, was more lenient than a statisical test using *Q*. Conventional heterogeneity tests estimate the mean effect size across several groups, and then assess or test, whether the mean effect size is consistent with all the studies. In contrast, when assessing clusterings of data with the model in Webster [32], equal means are assumed in all clusters, and minimising the log-likelihood determines whether the study data are best explained by one, or more clusters. This objective test is different to asking whether there are statistical differences between data, and is more tolerant of heterogeneity. It is easy to incorporate prior information into the approach, such as a weak normal prior for zero effect size. The selection of priors in a (1-dimensional) Bayesian meta-analysis is discussed in Ott et al. [34]. In principle, clustering methods could allow the identification of statistically distinct subgroups or outliers. This and the relative merits of the approach need considering in greater detail elsewhere, but it appears to offer a potentially useful alternative to conventional measures of heterogeneity.

### Composite endpoints in proportional hazards studies

Composite endpoints are frequently studied with proportional hazards models. This section explores the assumptions that are needed for the model to be correct, and for its estimates to be meaningful.

Firstly consider *m* diseases in a composite endpoint that may be influenced by the same factors or processes, but are otherwise independent of each other. The survival probability is determined from the probability of an individual surviving all *m* diseases until time *t*. If *S*_*j*_(*t*) is the probability of surviving disease *j* to time *t*, and *S*(*t*) is the probability of surviving all *m* independent diseases, then [15],

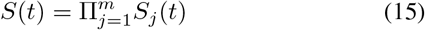

Writing *S*_*j*_(*t*) in terms of its hazard function *h*_*j*_(*t*), with 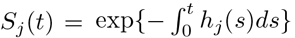, then Eq. 15 requires,

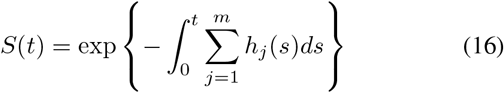

giving the hazard function *h*(*t*) for a group of diseases as [15],

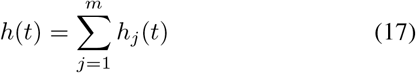

The proportional hazards model takes 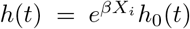 for an individual *i* with covariates *X*_*i*_, and assumes that *h*_0_ (*t*) is the same for all individuals in a population. If the assumption holds for each disease in a cluster or composite endpoint, then we can write,

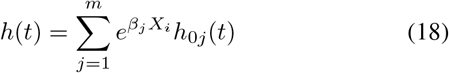

If all the diseases have the same risk-factor associations *β*, then,

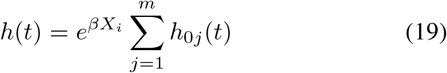

and the proportional hazards assumption will remain true for the cluster of diseases, with all the time-dependence in a single factor that will be the same for all individuals. However if any disease *k* has *β*_*k*_ *≠ β*, then,

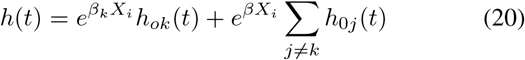

and the proportional hazards assumption fails. Note that this is the case if the assumption fails for any single covariate. The observations described by Eqs. 15-20 have the following implications:

1. For the proportional hazards assumption to hold, it is necessary that all clustered diseases have the same associations.
2. Counter-intuitively, the baseline hazard for each individual disease could be very different with diseases prevalent at very different ages, but the cluster of diseases can still satisfy the proportional hazards assumption.
3. If the proportional hazards model fails, it could be due to one or more of the clustered diseases having different risk associations - it need not necessarily be caused by different baseline hazards *h*_0*j*_ (*t*) in the underlying population.

Points 1 and 2 emphasise that diseases must be clustered on the basis of similar underlying etiology, and that incidence rates are irrelevant and potentially misleading for determining whether diseases can be considered as a single cluster and modelled with a proportional hazards model. Point 3 indicates that failure of the proportional hazards model, as identified by a statistical test using the Schoenfeld residuals for example [35], could be due to one or more diseases having different risk factors. This provides a consistency test for whether a cluster of diseases have the same risk factors, without fitting and comparing each disease individually, which may not always be possible.

### General remarks

If statistical tests are consistent with the proportional hazards assumption, then it can be reported that the diseases in the composite endpoint are consistent with having the same associations (within a proportional hazards model). This does not ensure that diseases have the same associations, only that there is no strong statistical evidence for them being different. Importantly, as was noted in the context of clinical trials, if the number of cases of a disease are few then they are unlikely to have much influence on a test result. In those cases the disease should only be included if there are strong prior (usually biological), reasons to include them.

The discussion above has considered the commonly used proportional hazard model. It has not considered the influence of model misspecification or the censoring distribution [36], or any newer methods for modelling composite endpoints that are being developed [37, 38].

### Multi-stage disease processes

Although Eq. 19 allows different baseline hazards for the diseases in a composite endpoint, the proportional hazards methodology requires that the resulting baseline hazard ∑ _*j*_ *h*_0*j*_ (*t*), is the same for all individuals (or individuals in a strata of a stratified analysis). Some rates of disease incidence, such as Cancers [16–18] and Amyotrophic Lateral Sclerosis [19–21], can be described by multi-stage disease processes [15], where one or more rate-limiting steps to disease may be skipped by a germline genetic variation. This will produce different baseline hazards for individuals with the genetic change. Importantly however, such changes will qualitatively modify the incident rate to a different power of time [15–21], and cannot be adjusted for in a proportional hazards model. Instead, the variant should be used to stratify the data, so that different strata have differing baseline hazards.

## Conclusions

Composite endpoints are intrinsic to how we define and study disease. Since John Graunt’s studies [5], there has always been a trade-off between disease definitions that are sufficiently specific to distinguish different underlying disease processes, but also sufficiently broad to allow a meaningful statistical study. This is particularly apparent in clinical trials and epidemiological studies where data are costly or unavailable. Large population datasets with detailed genetic and biological information are providing a new data-driven source of composite endpoints, by identifying composite endpoints with potential shared underlying causes.

Statistical methods can assess whether a composite endpoint is consistent with its assumed properties, such as testing its constituent diseases for heterogeneity among their disease-risk associations. Heterogeneity is conventionally tested with a *Q* or *I*^2^ statistic [12]. An alternative clustering-based approach, is to assume that diseases are in one or more clusters with equal associations, and test if the log-likelihood for the model [32] is minimised by one, or more clusters. This objective test can incorporate a prior, and examples suggest it is more lenient than a *Q* test, with the disease clustering data of Webster et al. [1] minimising the log-likelihood when *I*^2^ ≃ 50%. The merits of this approach for applications such as meta-analyses, will need exploring in greater detail elsewhere.

When proportional hazard models are used, then heterogeneity of associations in a composite endpoint will cause the proportional hazards assumption to fail. However, the statistical model does allow arbitrarily different baseline incidence rates for the diseases in the composite endpoint (although each baseline hazard *h*_0*j*_ must be the same for all individuals in the studied population). A test for the assumption of proportional hazards, provides a necessary consistency test when heterogeneity tests are impractical or impossible; although the proportional hazards assumption could fail for other reasons.

The multi-stage model of disease emphasises that germline genetic variants that modify the number of rate-limiting steps prior to disease, cannot be adjusted for within a proportional hazards model but must be used to stratify the data. This remark holds more generally, regardless of whether a disease is part of a composite endpoint or studied alone.

## Data Availability

UK Biobank data are available by application from www.ukbiobank.ac.uk. Simulated datasets used in the examples, will be made available via the Open
Science Foundation after publication.

## Appendix: Multivariate inverse variance weighted sum of squares

Note that the covariances and their inverses are symmetric, and expand,

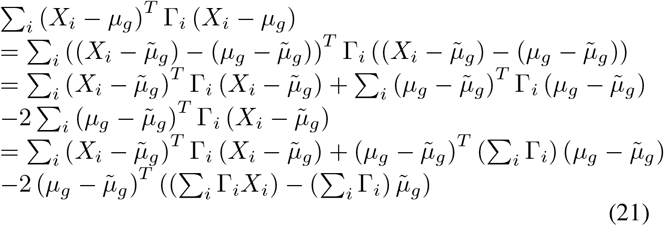

If 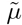 takes the specific form given by Eq. 6, then the terms (Σ_*i*_ Γ_*i*_*X*_*i*_) and 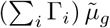 in the last term of the final line cancel, and the resulting equation can be rearranged to give,

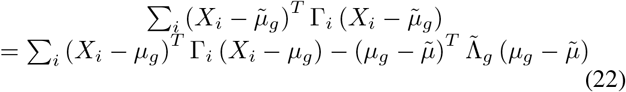

## Acknowledgements

This research has been conducted using data from UK Biobank, a major biomedical database, under application number 42583. Anthony Webster is supported by a fellowship from the Nuffield Department of Population Health (NDPH).

